# SARS-CoV-2 infection of BNT162b2(mRNA)-vaccinated individuals is not restricted to variants of concern or high-risk exposure environments

**DOI:** 10.1101/2021.05.19.21257237

**Authors:** Brittany Rife Magalis, Carla Mavian, Massimiliano Tagliamonte, Shannan N. Rich, Melanie Cash, Alberto Riva, Julia C. Loeb, Michael Norris, David Moraga Amador, Yanping Zhang, Jerne Shapiro, Petr Starostik, Simone Marini, Paul Myers, David Ostrov, John A. Lednicky, J. John Glenn Morris, Michael Lauzardo, Marco Salemi

## Abstract

The emergence of SARS-CoV-2 variants of concern (VOC) has raised questions regarding the extent of protection of currently implemented vaccines. Ten “vaccination breakthrough” infections were identified in Alachua County, Florida, among individuals fully vaccinated with the BNT162b2 mRNA vaccine as a result of social or household transmission. Eight individuals presented mild symptoms in the absence of infection with other common respiratory viruses, confirmed using viral genetic sequencing. SARS-CoV-2 genomes were successfully generated for five of the vaccine breakthroughs and 399 individuals in the surrounding area and were included for reference-based phylogenetic investigation. These five individuals were characterized by infection with both VOCs and low-frequency variants present within the surrounding population. Mutations, in the Spike glycoprotein, were consistent with their respective circulating lineages. However, we detected an additional mutation in Spike’s N-terminal domain of a B.1.1.7 strain, present at low-frequency (∼1%) in the unvaccinated population, potentially affecting protein’s stability and functionality. The findings highlight the critical need for continued testing and monitoring of infection among individuals regardless of vaccination status.

---

Several rapidly spreading variants of severe acute respiratory syndrome coronavirus 2 (SARS-CoV-2) have risen to the status of “variants of concern” (VOC), according to the Centers for Disease Control (CDC, https://www.cdc.gov/coronavirus/2019-ncov/cases-updates/variant-surveillance/variant-info.html), accumulating as many as 17 unique genomic mutations with respect to the original viral strain originating from the Wuhan province of China (1). These VOCs include the lineages most notably defined by their area of origin, such as the United Kingdom (UK). The UK VOC, also referred to as the B.1.1.7 lineage using the Pango nomenclature (2), emerged in September of 2020 in Kent, UK, but quickly made its way to the United States (US, (3)). More recently, the B.1.427/B.1.429 lineages, two forms collectively dubbed the “California variant”, were added to the CDC’s list of VOCs, as they have spread widely since their initial detection in July of 2020 in Los Angeles County, California (4). The emergence of these and other VOCs, and their level of evolutionary divergence from the original strain, has raised questions regarding the extent of protection of currently implemented vaccines against infection (5-8). Here, we report epidemiological and genomic data (where available) for vaccination breakthrough cases from Alachua County, Florida, in the context of the diverse SARS-CoV-2 epidemic in Florida and the remaining US.

## Results

From January to March 2021, Alachua County, Florida, experienced a reduced number of infections (∼11% drop in % positive cases), with a general increase in number of vaccinations, particularly since the last week of February (**Figure S1**). Regardless of the reduced coronavirus disease (COVID-19) burden in the county, continued testing of vaccinated individuals has provided insight into susceptibility of vaccines to circulating viral variants within the region. Between February and March 2021, ten individuals enrolled in the UF Health Screen, Test & Protect (STP) program (**online Methods**) tested positive using the standard polymerase chain reaction (PCR)-based assay for SARS-CoV-2 infection after completing the BNT162b2 mRNA two-dose vaccine series (9). Individuals were considered fully vaccinated two weeks after their second vaccination in a 2-dose series. By March 17, 2021, more than 59,000 individuals were vaccinated with at least one dose of one of Pfizer (49%), Moderna (49%), or Johnson & Johnson (2%) vaccines in Alachua County, Florida (10). The number of infections among vaccinated individuals suggests a relatively low breakthrough rate consistent with the high efficacy of Pfizer’s BNT162b2 vaccine demonstrated in clinical trials (9), though it is important to acknowledge the small sample size of this study and potential sampling biases incorporated in the STP program that limit drawing any conclusions regarding validation of efficacy (**online Methods**). The ten “vaccine-breakthrough” individuals were all identified within 27-47 (mean=37.6) days following the second dose, eight of which presented with symptoms characteristic of COVID-19 **(Tables 1** and **S1**). The majority of these individuals identified as female (90%), White (80%), under the age of 35 (mean=34), and reported their occupation as healthcare-related (80%), including students working in patient care. Contact tracing revealed that seven of these individuals had exposures to individuals previously identified as COVID-19 cases in the two weeks prior to their disease onset, with four reporting the nature of the relationship as household and three as community/social. None of the individuals reported exposure in the workplace. So, despite healthcare workers increased risk of exposure, infection in these individuals was likely the result of inadvertent exposure during social contact.

**Table 1.**
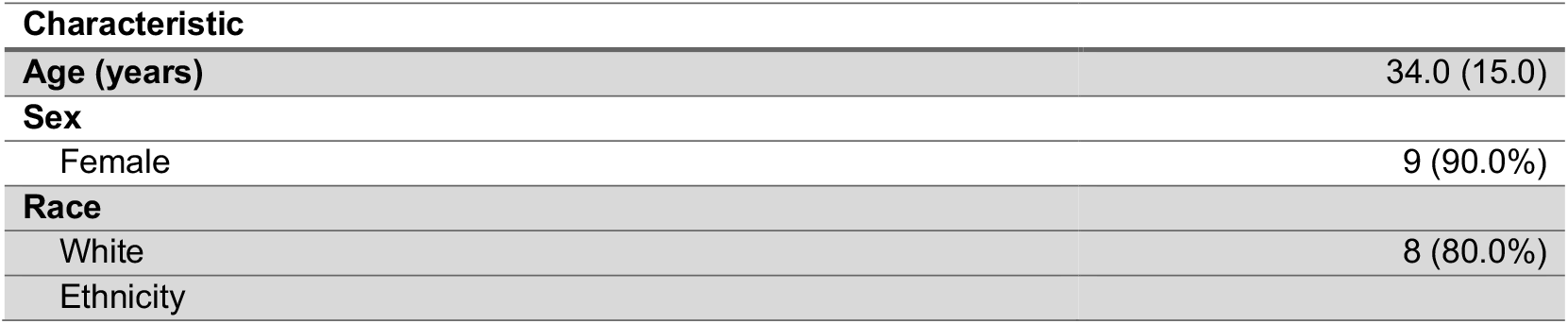
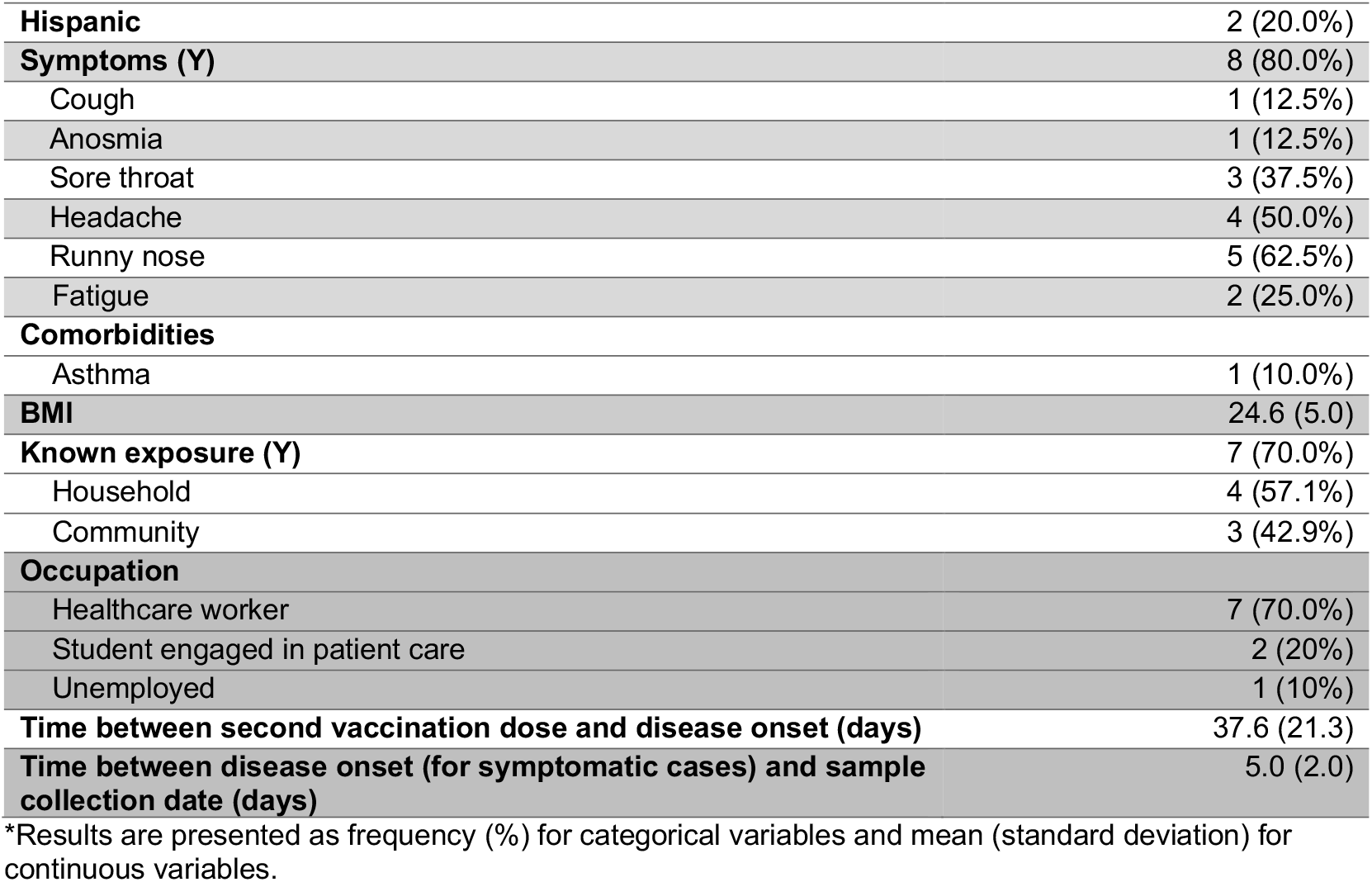
Reported cases of vaccination breakthroughs in Alachua County, Florida.

| Characteristic |  |
| --- | --- |
| Age (years) | 34.0 (15.0) |
| Sex |  |
| Female | 9 (90.0%) |
| Race |  |
| White | 8 (80.0%) |
| Ethnicity |  |
| **Hispanic** | 2 (20.0%) |
| **Symptoms (Y)** | 8 (80.0%) |
| Cough | 1 (12.5%) |
| Anosmia | 1 (12.5%) |
| Sore throat | 3 (37.5%) |
| Headache | 4 (50.0%) |
| Runny nose | 5 (62.5%) |
| Fatigue | 2 (25.0%) |
| **Comorbidities** |  |
| Asthma | 1 (10.0%) |
| **BMI** | 24.6 (5.0) |
| **Known exposure (Y)** | 7 (70.0%) |
| Household | 4 (57.1%) |
| Community | 3 (42.9%) |
| **Occupation** |  |
| Healthcare worker | 7 (70.0%) |
| Student engaged in patient care | 2 (20%) |
| Unemployed | 1 (10%) |
| **Time between second vaccination dose and disease onset (days)** | 37.6 (21.3) |
| **Time between disease onset (for symptomatic cases) and sample collection date (days)** | 5.0 (2.0) |

\*Results are presented as frequency (%) for categorical variables and mean (standard deviation) for continuous variables.

Secondary saliva samples were collected from the ten vaccine-breakthrough individuals within 3-7 days of testing positive for infection (**Tables 1** and **S1**). Saliva collection was used in this study for the isolation of viral RNA, as this bodily fluid has demonstrated prolonged presence of viral RNA (up to 25 days post-symptom onset, (11)), irrespective of disease severity (12). The research protocol (**online Methods**) was approved by the University of Florida institutional review board. SARS-CoV-2 full-genome (>70%) sequences were successfully generated for five of the vaccinated individuals. A panel of additional respiratory viruses was also targeted during sequencing (**Table S2**), confirming the absence in all patients of additional infection with common viruses, such as influenza. Thus, clinical symptoms, when present, were likely the result of productive coronavirus infection. The reason for insufficient SARS-CoV-2 genome quality for five of the vaccine breakthrough cases is not fully clear. Results from COVID-19 testing for the majority of these individuals were limited to qualitative data (positive or negative), though the number of PCR cycles (Cq) required for reliable viral RNA amplification for two of the individuals were provided and already >25 at the time of diagnosis (**Table S3**), which has been considered relatively high for genomic sequencing (13). Given the time between diagnosis and saliva sampling (**Table S1**), saliva viral load may have been too low for the genomic amplification required for amplicon-based sequencing for some individuals. While mutations in primer-binding regions can reduce genome coverage for amplicon-based sequencing (13), coverage mapping did not indicate this phenomenon – alternating regions of high- and low-coverage were not observed (**Figure S2**). Two of these individuals were, however, asymptomatic whereas asymptomatic individuals were not found among the successful full-genome sequences (**Table S2**), supporting the link between severity of symptoms and rapidity of viral clearance (14).

In order to understand the context of the vaccine breakthrough infections, we assembled a full-genome sequence dataset (n=3902) including sequences generated from hospital samples in Alachua County between January-March 2021 (n=399), all Floridian sequences (n=3,016) deposited into the GISAID database (https://gisaid.org), and epidemiologically relevant sequences from the global population (also from GISAID). Epidemiological relevance was defined on an individual sequence basis, restricting the global GISAID search to the two sequences most similar genetically to each Floridian sequence and sampled within a high-confidence transmission time window (30 days) based on sample collection date. Additional details on this method (termed “FLACO-BLAST”) can be found in the **online Methods** and (15), and the script is available from https://github.com/salemilab/flaco. GISAID IDs and corresponding submission information are provided in **Table S4**. Owing to low genetic variability, as well as potentially shared epidemiological linkages, sequences retrieved from GISAID were often shared by more than one query (Florida) sequence, resulting in a total of 482 non-Floridian sequences, all of which were located within the US. The resulting dataset spanned October 10, 2020 to March 17, 2021.

Lineages for all sequences were determined using the PangoLEARN model (Pangolin v 2.3.6), which was trained using ∼60,000 GISAID SARS-CoV-2 sequences to classify incoming sequences based on molecular and epidemiological criteria (2). The full dataset in this study was characterized by a total of 106 lineages, for which only nine (B.1.X) lineages represented >99% of samples (**Figure 1A**). The distribution of lineages for Floridian sequences outside of Alachua County largely resembled that of the non-Floridian reference sequences, as expected given the filtering approach for genetic similarity described above; the exception to this similarity was the presence of the B.1.375 lineage within the Florida dataset, which was not present among reference sequences (**Figure 1A**). As this lineage was not observed among the Alachua County sequences, further investigation into the potential misclassification of these sequences as the B.1.375 lineage, more notably associated with the northern states (16), was considered outside the scope of this study. Whereas both the Alachua County and remaining Floridian samples were dominated by the B.1.234 lineage in November 2020, both regions quickly expanded to include at least seven other lineages by mid-January 2021 (**Figure 1A**). This expansion included a growing presence of the B.1.1.7 (or UK VOC) and B.1.427 (California VOC) lineages (**Figure 1A**). Given the growth in the UK and California variants within the Florida population, it is not surprising that three of the vaccinated individuals reported in this study were determined to belong to these VOCs – 1 B.1.1.7 and 2 B.1.247/B.1.249. The remaining two vaccinated individuals, however, presented with the high-frequency (∼16%) B.1 lineage and low-frequency (<1%) B.1.377 lineage, neither of which is considered a VOC.

**Figure 1.**
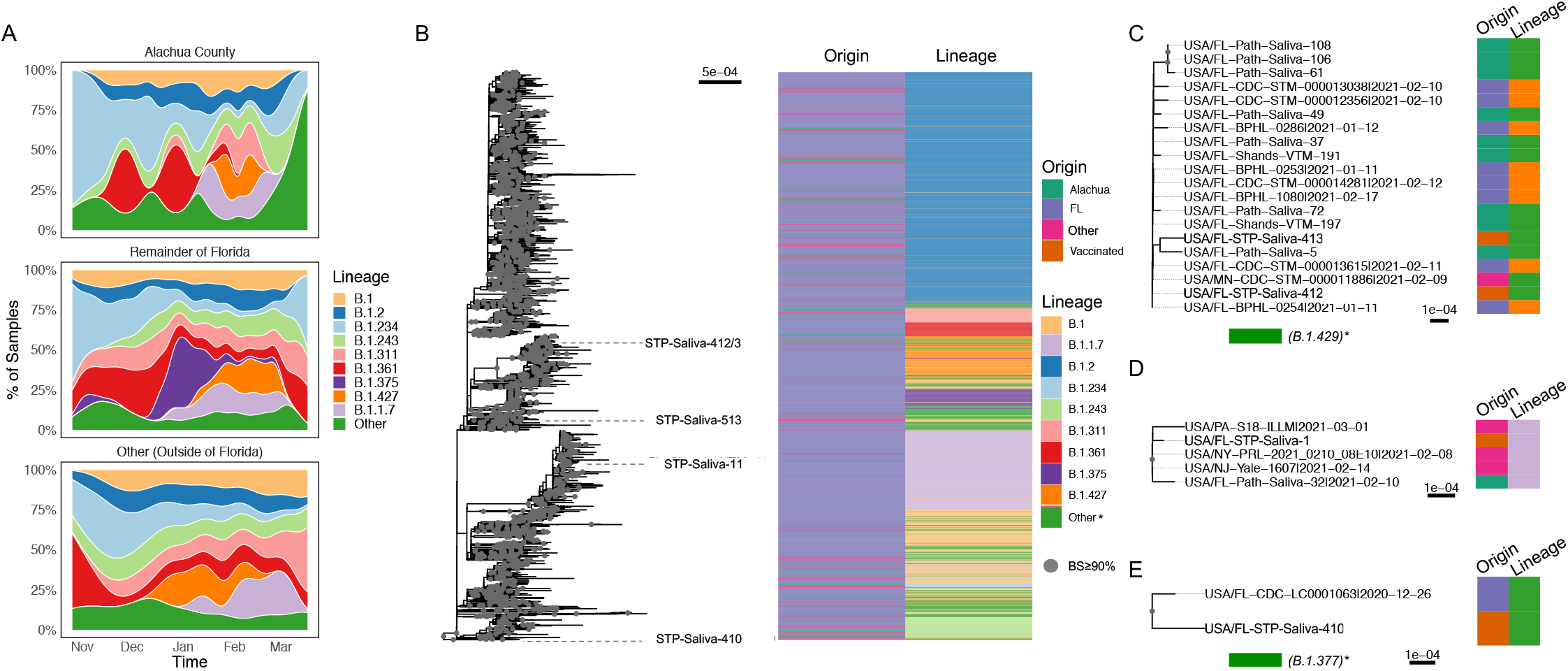
Distribution of identified lineages and geographical origin of sampling over time (A) and within the phylogenetic tree (B-D) of SARS-CoV-2 data collected from Florida and relevant non-Florida locations. (A) Lineage (as identified using Pangolin (2) distribution over time for Alachua County, the surrounding Florida areas, and locations outside of Florida linked to Florida sequences via genetic similarity. (B) Distribution of both assigned lineages and geographic origin (as in (A)) across the maximum likelihood phylogenetic tree. Branches are scaled in genetic substitutions/site, and nodes with ≥90% support using bootstrap sampling are indicated by grey dots. Vaccinated individuals within well-supported clades have been emphasized, and corresponding clades are represented as insets in panels C-D. *Other lineages, defined as present within <1% of the total sample population.

Following lineage classification, a maximum likelihood phylogenetic tree was reconstructed from the sequences in order to verify lineage classification and to determine relationships among vaccinated individuals in the context of geographical and temporal information (**Figure 1B**). Bootstrap replicates for the sequence data were used to provide support for branching patterns within the tree (17-19), and the smallest (number of sequences), well-supported (>90% of replicates) clade involving each vaccination-breakthrough case was examined individually. Whereas four of the five successfully sequenced, vaccinated individuals belonged to relatively small, definable clades (**Figure 1C-D**), reliable placement of the fifth B.1-lineage individual within the tree could not be obtained, despite >90% coverage of the genome (**Figure S2**). This sequence did, however, share common ancestry with similarly B.1 sequences, supporting proper lineage assignment (**Figure S3**). The two individuals harboring the B.1.429 California VOC belonged to a clade of 20 individuals comprised of additional B.1.429/B.1.427 variants, confirming lineage classification using Pangolin (**Figure 1C**). The remaining 18 sequences within this clade consisted primarily of Floridian sequences (nine from Alachua County), with the exception of one individual from Minnesota, suggesting largely local transmission of this variant, though directionality of transmission cannot be inferred. The two vaccine-breakthrough cases with the B.1.429 VOC in this study (STP-413 and STP-412) reported separate exposures; the two individuals were also not more closely related to each other phylogenetically than the remainder of the sequences (with significant support), so we can neither confirm nor exclude the possibility of a relationship via direct transmission between these two individuals specifically.

Circulation of the other VOC in the US, B.1.1.7, has been defined as the result of at least two separate introductions (3). One introduction was estimated to occur early November, 2020, into California, and was characterized by a virus resembling more closely the traditional UK variant. The second introduction, estimated to occur in late November, carried a silent mutation from cytosine to thymine within the ORF1ab gene encoding the RNA-dependent RNA polymerase (position 15720 relative to genome start). The expanding virus populations originating from these two events were referred to as “Clade 1” and “Clade 2.” Approximately 13% of B.1.1.7 individuals within the total Florida sample, and 86% of B.1.1.7 in Alachua County, harbored the “Clade 2” C15720T mutation, including the vaccine-breakthrough case. This individual (STP-11) belonged to a well-supported clade in this study comprised of five total individuals (**Figure 1D**), all sharing the B.1.1.7 lineage designation (again confirming pangolin classification) and “Clade 2” mutation. Despite the prevalence of the “Clade 2” mutation among B.1.1.7 individuals in Alachua County, only one other sample within the vaccine-breakthrough clade originated from the county. The remaining four samples were collected in Pennsylvania, New York, and New Jersey, consistent with the earlier evidence of a more widespread presence of this B.1.1.7 variant within the US (3). The individual was reportedly exposed to a recent COVID-19 case outside of the household. Contact tracing of this individuals’ exposed contacts did not identify any secondary cases associated with this vaccine failure, and so the question of ongoing transmission following infection in this individual remains unanswered.

The final clade containing the low-frequency B.1.377 vaccinated individual (STP-410) shared significant common ancestry with a single Floridian B.1.377 sequence, confirming lineage classification and suggesting transmission within Florida, but not confined to the county (**Figure 1E**).

Even though infection in fully vaccinated individuals does not appear to be related to the viral variant, mutational analysis was necessary to determine if 1) recently acquired mutations (i.e., not conserved among the corresponding lineage) could potentially be responsible for limited protection of the vaccine against infection or 2) evidence exists for the emergence of vaccine-resistant variants. The viral protein commonly referred to as the Spike protein is structurally important for coronaviruses, rendering it susceptible to recognition by the host immune response, specifically the receptor-binding domain (RBD). For this reason, vaccination efforts have largely focused on this region of the genome (9, 20, 21), though there is evidence that the host immune system can also target the N-terminal domain (NTD, (22, 23)). The natural accumulation of mutations in these regions owing to the error-prone viral replication machinery can lead to the ability of the virus to evade the host immune response, including that of vaccinated individuals, and may be responsible for enhanced transmissibility of the variant (24). The B.1.429 and B.1.377 variants found within three of the breakthrough-vaccinated individuals did not appear to have acquired any additional Spike mutations outside of those associated with the parental lineage, suggesting newly acquired mutations were not responsible for breakthrough infections and that this individual did not harbor a vaccine-resistant variant. However, two mutations relative to the parental B.1.1.7 lineage (1) were observed within the Spike protein for the B.1.1.7 vaccinated individual – L18F and K1191N within the NTD and C-terminal domain (CTD) of the Spike protein, respectively (**Figure 2A**, corresponding to positions within the reference sequence MN908947). The HR2 K1191N mutation was present in ∼75% of samples for Alachua County, ∼75% of the remainder of Florida samples, and ∼75% of US reference sequences, suggesting a less recent acquisition of this mutation in the population and no relation to vaccine-mediated adaptation or an enhanced ability to infect vaccinated individuals. Alternatively, the NTD L18F mutation was far less prevalent, only observed in 16 individuals from Florida (dating back to January 15, 2021), but not from the Alachua County area (**Figure 2B**). It was known that the B.1.1.7 vaccination-breakthrough individual (STP-11) was exposed to an infected individual from Orange County, Florida, which suggests a regional Florida outbreak with this particular mutation that spilled over into Alachua County. No evidence of secondary cases associated with this individual again cannot confirm ongoing transmission of this variant among vaccinated individuals, though this mutation may warrant further investigation for its antigenic properties and relationship to vaccination efficacy.

**Figure 2.**
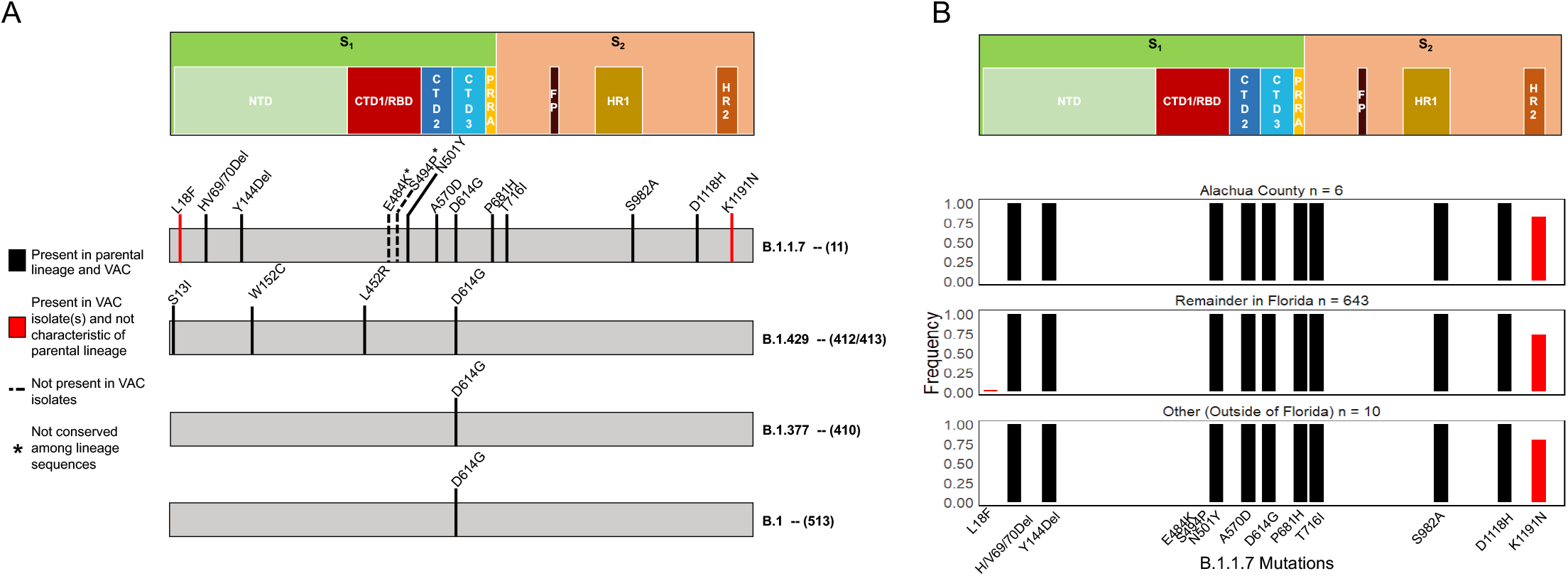
Mutational profiles for vaccine-breakthrough individuals (A) and comparison of B.1.1.7 individuals with remaining sequence data (B). Mutations in red are present in vaccinated individual (VAC), but not fixed in the parental lineage. Spike protein architecture is displayed at the top of both panels, wherein S = spike subunit (1-2); NTD = N-terminal domain; CTD (1-3) = C-terminal domain; RBD = receptor binding domain; PRRA = SARS-CoV-2 characteristic PRRA insertion at the S_1_/S_2_ cleavage site; FP = fusion peptide; HR (1-2) heptad repeat. Positions numbers are relative to the Spike protein in the MN908947 reference sequence.

Several molecular vaccine and/or antigenic studies in the context of the B.1.1.7 variant have focused on the RBD, demonstrating no significant effect on neutralization of the virus (25) and no loss of affinity for host antibodies (Abs) to the RBD when compared to the original strain. Meanwhile, mutations in the NTD (other than L18F) have been proposed to alter Ab affinity, potentially paving the way for infection in vaccinated individuals (20). The structural consequence of the recently acquired NTD L18F mutation indicates that this Floridian B.1.1.7 variant may be a variant of concern. In addition to B.1.1.7 deletions that eliminate antigenic epitopes for neutralizing antibodies (26) and T cells (Δ69-70, Δ144), this variant exhibits an amino acid change with the potential to stabilize the NTD of the Spike protein. Amino acid position 18 is located within the N1 loop (residues 14 to 26), wherein the original L (Leucine) is oriented towards F (Phenylalanine) at position 79 of the N2 loop (residues 67 to 79, **Figure 3A**). The change from L to F in the Florida B.1.1.7 variant is predicted to form stabilizing contacts between F18 in the N1 loop and S252 in the N5 loop (residues 246 to 260) (**Figure 3B**). These data suggest that the Floridian B.1.1.7 variant found in the B.1.1.7 vaccinated individual may exhibit a fitness advantage that results from increased stability and functionality of the NTD.

**Figure 3.**
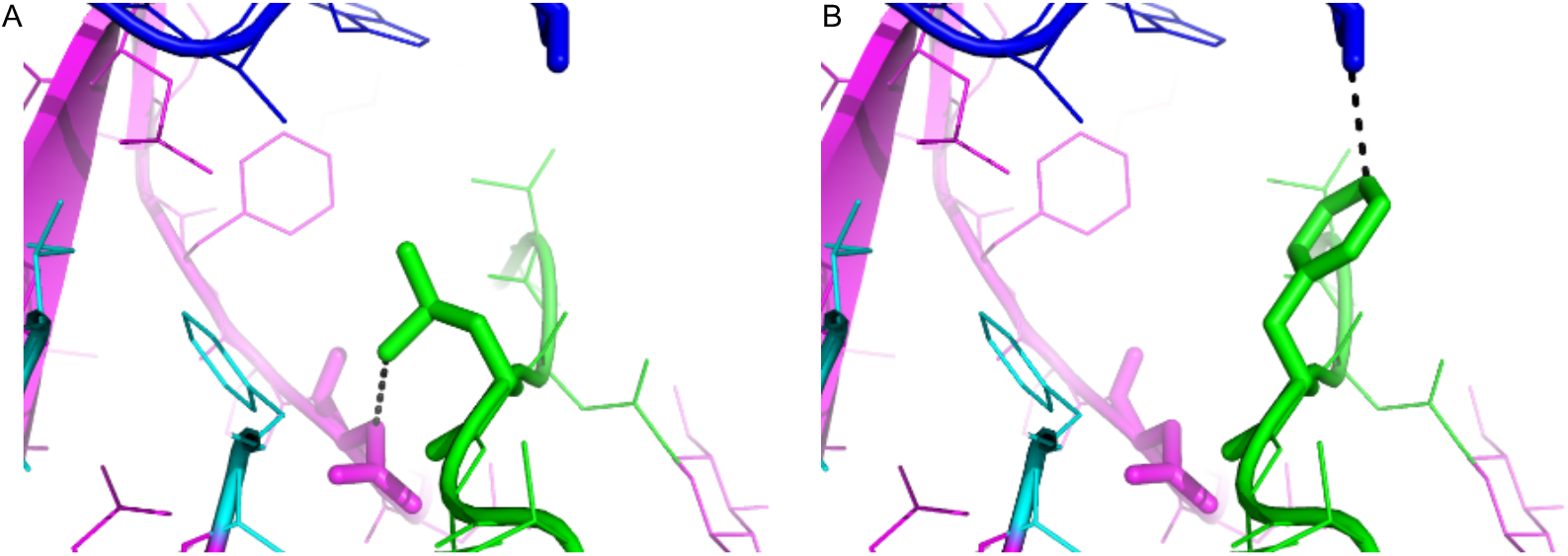
Potential stabilizing effect of Floridian B.1.1.7 variant mutation from Leucine (A) to Phenylalanine (B) at position 18 of the Spike protein N-terminal domain (NTD). Loops N1, N2, and N5 of the NTD are represented in green, pink, and blue, respectively. Modified interactions as a result of the mutation are represented as dotted lines, with original L18 oriented toward position F79 (cyan) of the N1 loop (A), and the variant F18 toward S252 of the N5 loop (B), potentially acting to stabilize. Dotted lines represent distances 3.8 (A) and 3.6 (B) Angstrom. Positions numbers are relative to the Spike protein in the MN908947 reference sequence.

In order to verify viability of this new mutation, infectivity of the virus isolated from individual STP-11 was measured *in vitro* (**online Methods**). Positive infection, and thus viability, was defined by the presence of virus-specific cytopathic effects (CPE) on Vero E6 cells (African green monkey kidney cells). CPE were first noted 12 days post-inoculation (dpi), primarily consisting of rounding of cells, some in the process of detaching from the growing surface (**Figure 4**). Quantification of virus at 12 dpi for the STP-11 sample-infected cells and control (no viral inoculation) confirmed viral replication (C_q_ 8.14 and C_q_ > 39, respectively).

**Figure 4.**
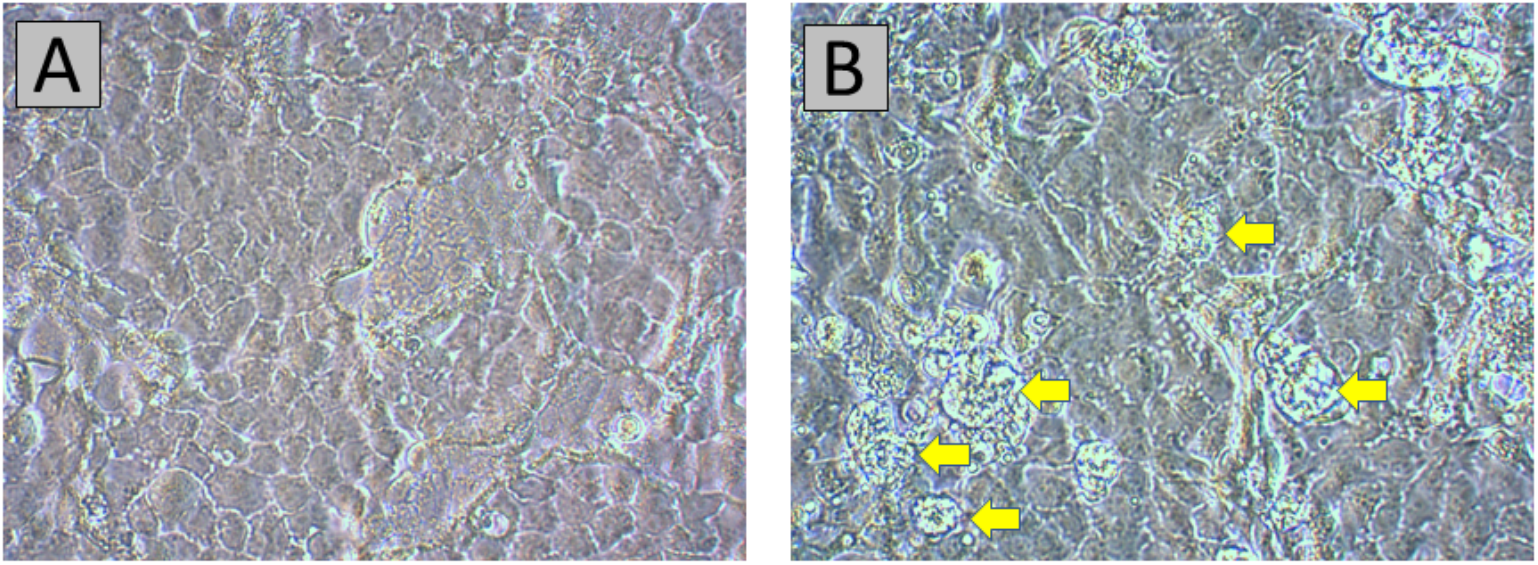
Cytopathic effects in Vero E6 cells inoculated with saliva sample FL-STP-Saliva-11. (A) Mock-infected Vero E6 cells, 12 dpi. (B) Early SARS-CoV-2-specific CPE, 12 dpi. Rounded cells, some in the process of detaching from the growth surface, are pointed out by yellow arrows. Original magnification at 400X.

## Discussion

A recent study in Israel reported that vaccinated individuals were disproportionally infected with VOCs relative to the unvaccinated population (27). Whereas we cannot exclude the impact of prevalence of VOCs within the Florida population on their rate of breakthrough, the findings of our study, collectively with (27), indicate limited protection of the BNT162b2 mRNA vaccine (and potentially others) against emerging variants of SARS-CoV-2. It is important to note that none of the breakthrough individuals in this study was hospitalized, corroborating the vaccine’s 100% efficacy against severe disease (9). While these individuals presented with only mild symptoms (or no symptoms at all), the number of vaccine breakthrough cases might be expected to be under-reported. Hence, if we assume at least a minority of test-positive vaccinated individuals harbor infectious virus, the potential for hidden reservoirs within the global populations is increased. Hidden reservoirs in asymptomatic or mild symptomatic individuals, as has been proposed (15), pose a particular threat to early recognition of mutations of potential concern, such as the L18F mutation described herein, particularly once vaccination is more widespread. Moreover, given the effectiveness of the vaccine in limiting symptom presentation, vaccinated individuals may engage more frequently in social activities, increasing the risk of exposure. Continued testing and case management, assessing contacts and exposure, for vaccinated individuals is thus encouraged and will be forthcoming in determining whether the vaccine is protective against ongoing evolution and spread of SARS-CoV-2. This strategy is particularly important in the face of relaxed guidelines regarding masked protection and social distancing for vaccinated individuals, declared by the CDC as of May, 2021.

## Data Availability

GISAID sequences used for the study are described in the Supplementary Material. Newly added sequences will be submitted to GISAID following acceptance but are available from the authors (along with sequence quality information) until such time.

## Acknowledgments

This work was supported by the Stephany W. Holloway University of Florida Chair and by funds of the University of Florida Office of Research and Health Science Center with resources from the Interdisciplinary Center for Biotechnology Research Gene Expression Core (RRID:SCR_019145), NextGen Sequencing Core (RRID:SCR_019152) and Bioinformatics Core (RRID:SCR_019120). Funding for this work was also provided by National Science Foundation (NSF) – Division Of Environmental Biology (DEB) award no. 2028221.

## Methods

### Participant involvement and sample processing

UF Health Screen, Test & Protect (STP) assists the Florida Department of Health in Alachua County with COVID-19 case and contact tracing efforts in its UF students, faculty, staff and other UF-affiliated people including the UF Health Academic Medical Center (∼123,000 total UF Affiliates). Full epidemiological investigations were conducted on positive cases to collect exposure information, trace contacts, and provide disease transmission education. Fully vaccinated individuals who became a contact (defined as ≥15min and closer than 6 feet) were called and provided public health education but not placed into quarantine. For 14 days after their last exposure, they received a daily text or email to record symptom development. Immediate PCR testing was recommended for anyone newly reporting symptoms and day 7 PCR testing was recommended for all fully vaccinated but exposed individuals, regardless of symptom development. Individuals working in health care settings may have been sampled more frequently due to internal hospital policies.

Fully vaccinated UF affiliates deemed PCR-positive for SARS-CoV-2 were eligible for molecular epidemiology investigation as part of the STP program if they met the following criteria:

1. The case must be infectious at the time of saliva sample donation. Infectiousness was defined ≤10 days after the onset of symptoms or for an asymptomatic individual ≤10 days after the positive lab collection date.
2. The case must meet the definition of a vaccine-breakthrough case. Defined as PCR-positive for SARS-CoV-2 and ≥14 days after the second dose of Pfizer or Moderna.

If both criteria were met and individuals volunteered to provide a sample for the purpose of public health molecular surveillance, they were scheduled to arrive on-site for sample collection as soon as possible to increase the probability of detectable virus at the time of collection. Each participant was asked to give a saliva sample of at least 2 mL in total volume and was instructed not to drink anything for 10 minutes prior to giving the sample. The saliva was collected in a 15 mL conical tube, filling it to the 2 mL marking on the tube, not including froth. Patient samples were de-identified following Institutional Review Board approval before viral processing. Viral RNA was extracted from 180 uL of each saliva sample using the QIAamp 96 Viral RNA Kit with the QIAcube HT (Qiagen, Germantown, MD) using the following settings with a filter plate: the lysed sample was premixed 8 times before subjecting to vacuum for 5 minutes at 25kP and vacuum for 3 min at 70kPa. Following 3 washes using the same vacuum conditions above, the samples were eluted in 100 uL AVE buffer followed by a final vacuum for 6 minutes at 60kPa. Nine microliters of RNA was used for cDNA synthesis and library preparation using the COVIDSeq Test kit (Illumina, San Diego, CA) and Mosquito HV Genomics Liquid Handler (SPT Labtech Inc., Boston MA). The size and purity of the library was determined using the 4200 TapeStation System (Agilent, Santa Clara, CA) and the Qubit dsDNA HS Assay Kit (Life Technologies, Carlsbad, CA) according to the manufacturer’s instructions. Constructed libraries were pooled and sequenced using the NovaSeq 6000 Sequencing System SP Reagent Kit and the NovaSeq Xp 2-Lane Kit. Illumina’s DRAGEN pipeline was used to derive sample consensus sequences, which were filtered based on a minimum of 70% coverage of the genome.

### Database sequence retrieval

Each Floridian sequence was used in a local alignment (BLAST) (28) search for the most (genetically) similar non-Floridian sequence in the GISAID database as of March 23, 2021, and linked to two reference sequences including the best match (highest E-value) with a date occurring within one month following, as well as one month prior to the sampling date of the Floridian sequence (15). After removing duplicate sequences (sequences with same GISAID ID), sequences were aligned in viralMSA (29) using the MN908947 reference sequence, and mutations potentially associated with contamination, recurrent sequencing errors, or hypermutability were masked using a vcf filter (https://virological.org/t/masking-strategies-for-sars-cov-2-alignments/480).

### Cell culture for viral infectivity

Vero E6 cells were used for SARS-CoV-2 isolation attempts. The cells had been obtained from the American Type Culture Collection (catalog no. ATCC CRL-1586) and have been used for our SARS-CoV-2 projects (30-33), including the isolation of > 30 SARS-CoV-2 isolates from human and environmental samples. The cells were propagated in cell culture medium comprised of aDMEM (advanced Dulbecco’s modified essential medium, Invitrogen, Carlsbad, CA) supplemented with 10% low antibody, heat-inactivated, gamma-irradiated fetal bovine serum (FBS, Hyclone, GE Healthcare Life Sciences, Pittsburgh, PA), L-alanine, L-glutamine dipeptide supplement (GlutaMAX,), and 50 μg/mL penicillin, 50 μg/mL streptomycin, 100 μg/mL neomycin (PSN antibiotics, Invitrogen) with incubation at 37°C in 5% CO_2_.

### Isolation of virus in cultured cells

Virus isolation attempts were performed in a BSL3 laboratory at the University of Florida Emerging Pathogens Institute (EPI) by analysts who wore powered air-purifying respirators and engaged in BSL3 work practices. Vero E6 cells grown as monolayers in a T-25 flask (growing surface 25 cm^2^) were inoculated when they were at 80% of confluency as follows: for each flask, the spent cell culture medium was removed and replaced with 1 mL of supplemented aDMEM medium (“complete medium”) with 10% FBS, and the cells inoculated with 50 μL of unfiltered saliva. Prior to inoculation, samples were frozen (−80°C) within 15 minutes following collection. Samples were only thawed once to produce an aliquot for processing. The inoculated cell cultures were incubated at 37°C in 5% CO_2_, and rocked every 15 minutes for 1 hour, after which 4 mL of complete medium with 10% FBS was added. The following day, the cell culture media was completely removed and replenished with 5 mL of maintenance medium (complete medium with 3% FBS). Mock-infected cell cultures were maintained in parallel with the other cultures. The cell cultures were refed every 3 days by the replacement of 2 mL of spent media with maintenance medium. The cells were observed daily for one month before being judged negative for virus isolation, with a blind passage performed 15 days post-inoculation of the cells. When virus-induced cytopathic effects (CPE) were evident, the presence of SARS-CoV-2 in the cell culture medium was examined by real-time reverse transcription PCR (rRT-PCR). In the event that SARS-CoV-2 strains that were not cytolytic or did not produce CPE had been isolated, the culture media were blindly tested at weekly intervals.

### Detection of SARS-CoV-2 genomic RNA (vRNA) in cell culture medium

vRNA was extracted from virions in collection media in a Class II biosafety cabinet in a BSL3 laboratory at the EPI by analysts wearing appropriate personal protective equipment (chemically impervious Tyvek lab coats and gloves) and using powered-air purifying respirators. The vRNA was extracted from 140 µL aliquots of the collection media using a QIAamp Viral RNA Mini Kit (Qiagen, Valencia, CA, USA), and purified RNA eluted from the RNA-binding silicon column in a volume of 80 µL. Twenty-five µL (final volume) rRT-PCR tests were performed in a BioRad CFX96 Touch Real-Time PCR Detection System using 5 µL of purified vRNA and the N1 and N2 primers and their corresponding probes of the CDC 2019-Novel Coronavirus (2019-nCoV) rtRT-PCR test (34). The primers and probes were purchased from Integrated DNA Technologies (IDT, Coralville, Iowa, USA). A plasmid that encodes the SARS-CoV-2 N-gene sequence was purchased from IDT and used in positive control reactions for the CDC N1 and N2.

The rRT-PCR tests were performed using the following parameters: 400 nM final concentration of forward and reverse primers and 100 nM final concentration of probe using a SuperScript III One-Step RT-PCR system with Platinum Taq DNA Polymerase (ThermoFisher Scientific). Cycling conditions were 20 minutes at 50°C for reverse transcription, followed by 2 minutes at 95°C for Taq polymerase activation, then 45 cycles of denaturation for 15 seconds at 95°C, annealing for 30 seconds at 55°C, and extension at 68°C for 20 seconds.

**Table S1.**
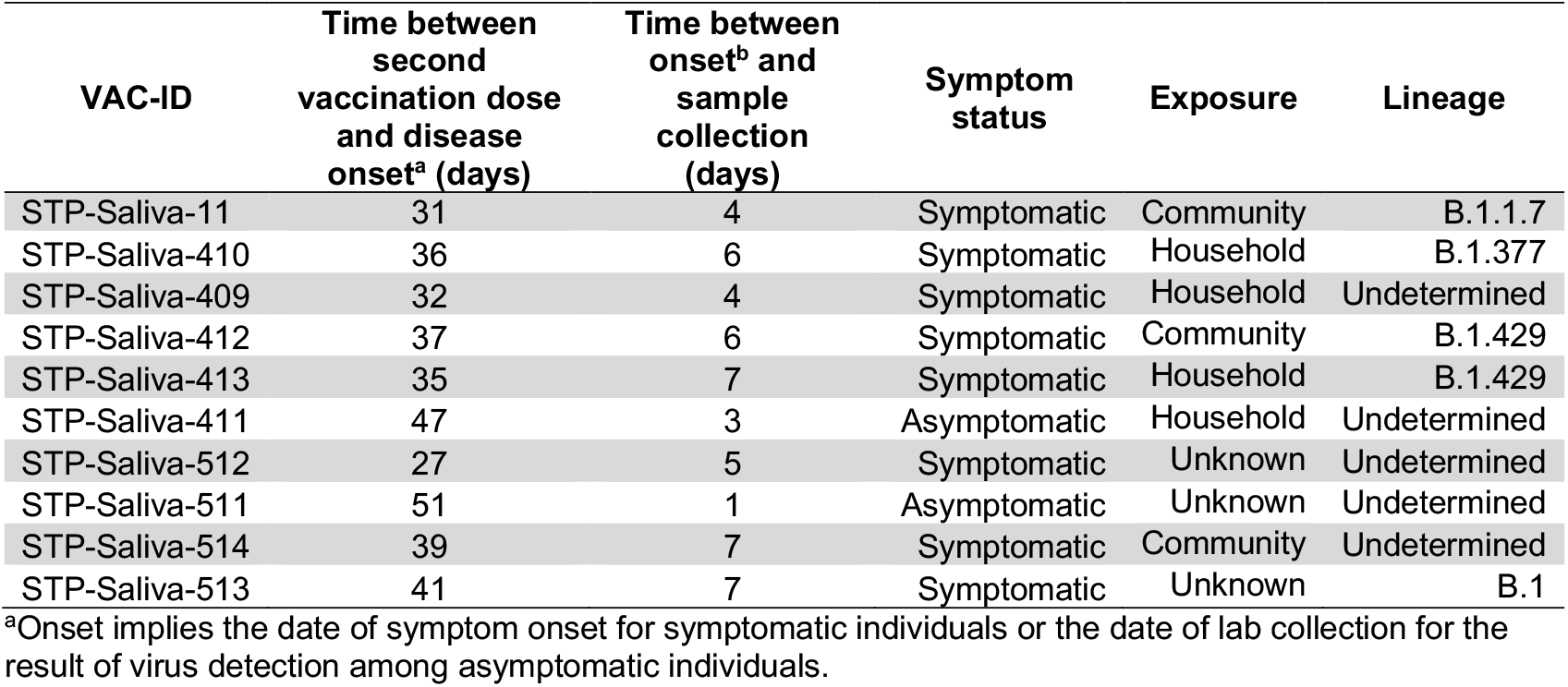
Sample and sequencing information for vaccinated (VAC) individuals.

**Table S2.**
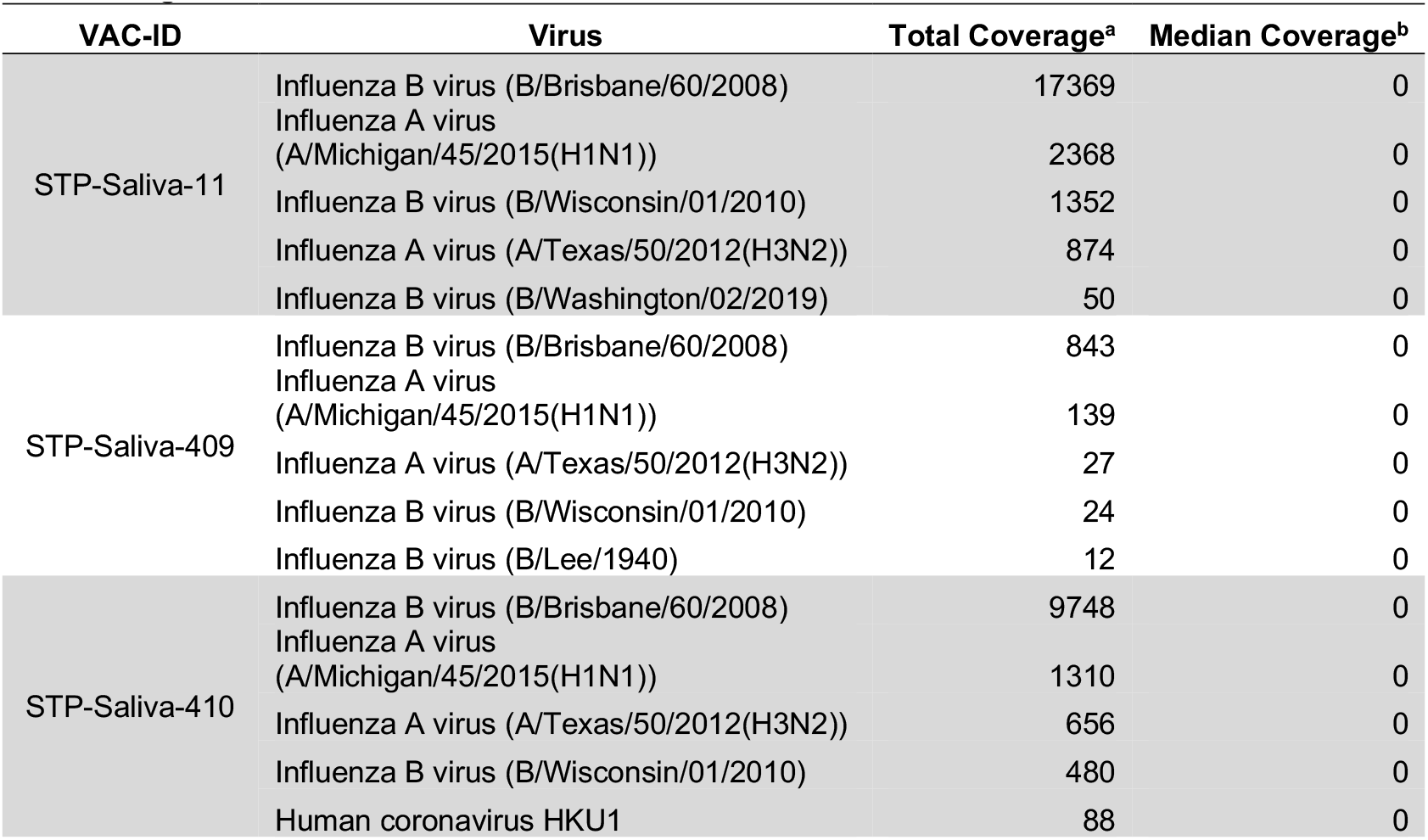
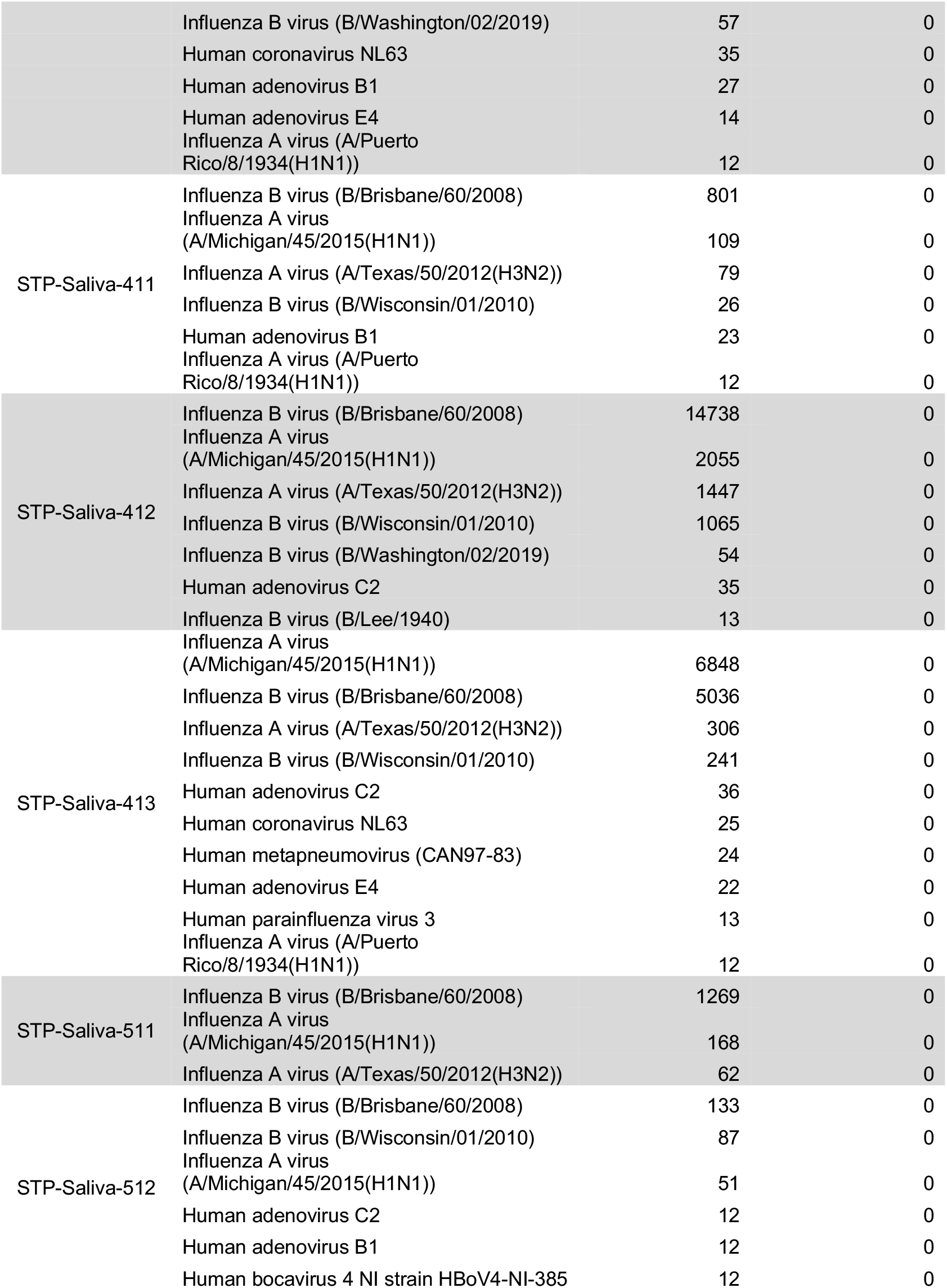
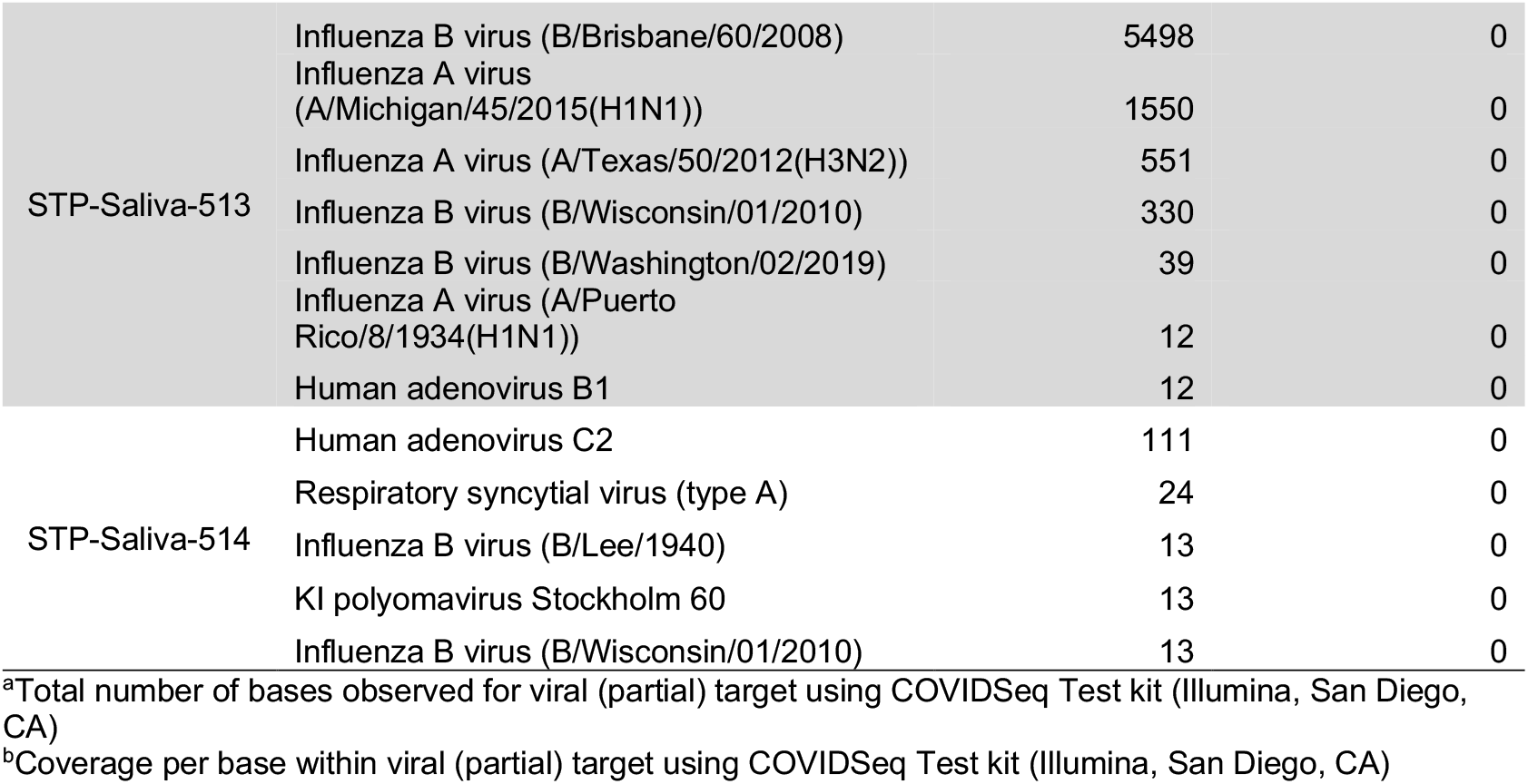
Respiratory virus coverage during sequencing of saliva samples from vaccine-breakthrough individuals.

**Table S3.** Quantification results from initial detection of SARS-CoV-2 (at time of diatnosis) in vaccinated individuals.

| VAC-ID | Cq<sup>ab</sup> (Time of diagnosis \| Time of secondary saliva sample collection) |
| --- | --- |
| STP-Saliva-11<sup>b</sup> | Not available |
| STP-Saliva-410<sup>b</sup> | Not available |
| STP-Saliva-409<sup>b</sup> | Not available |
| STP-Saliva-412<sup>b</sup> | Not available |
| STP-Saliva-413<sup>b</sup> | Not available |
| STP-Saliva-513<sup>b</sup> | Not available |
| STP-Saliva-411 | Not available |
| STP-Saliva-512 | Not available |
| STP-Saliva-511 | 36.48 \| 30.74 |
| STP-Saliva-514 | 28.85 \| 35.59 |

<sup>a</sup>PCR quantification cycle
<sup>c</sup>Vaccinated individuals with 70% genome coverage at 50x sequencing coverage/site.
\*

**Table S2.**
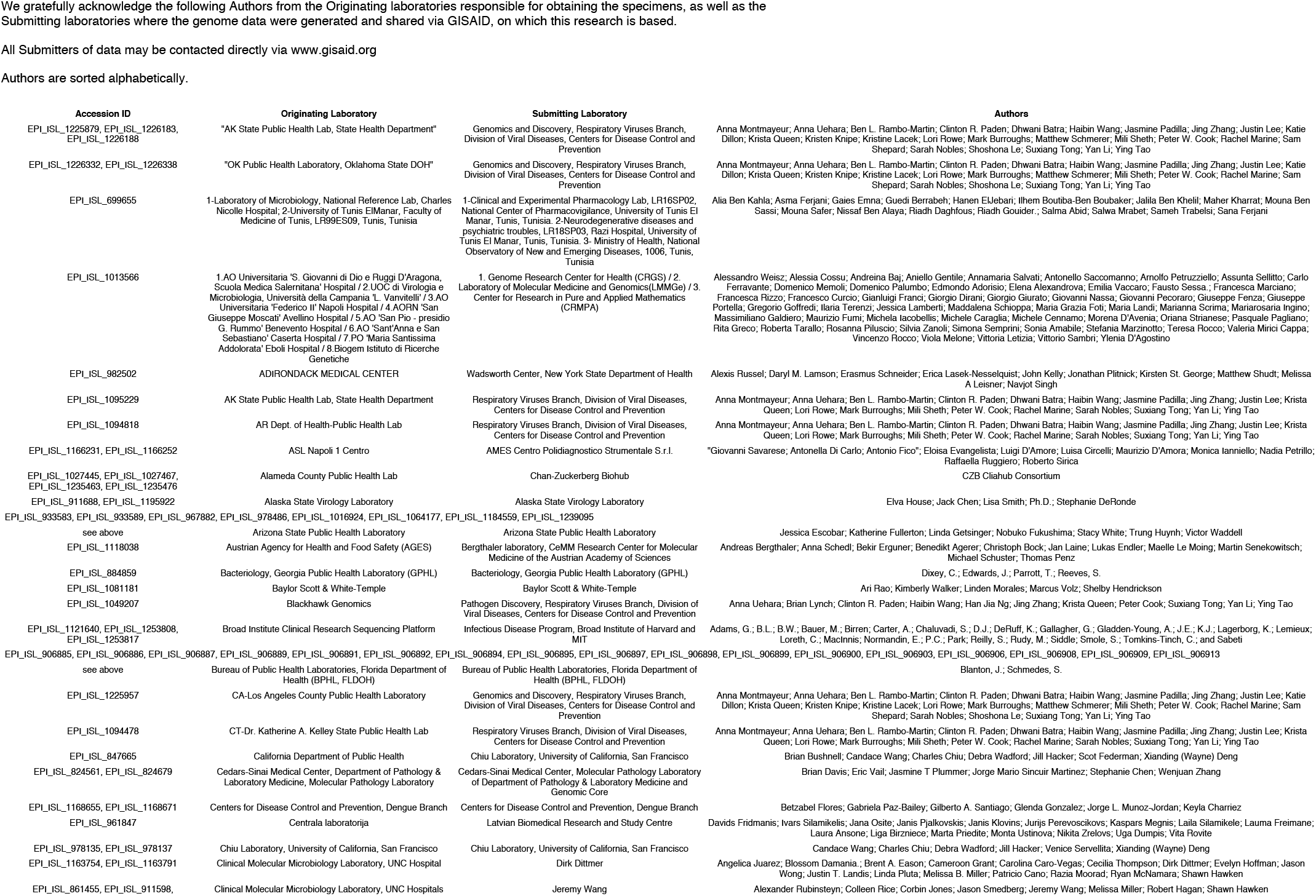
GISAID author acknowledgment table.

**Figure S1.**
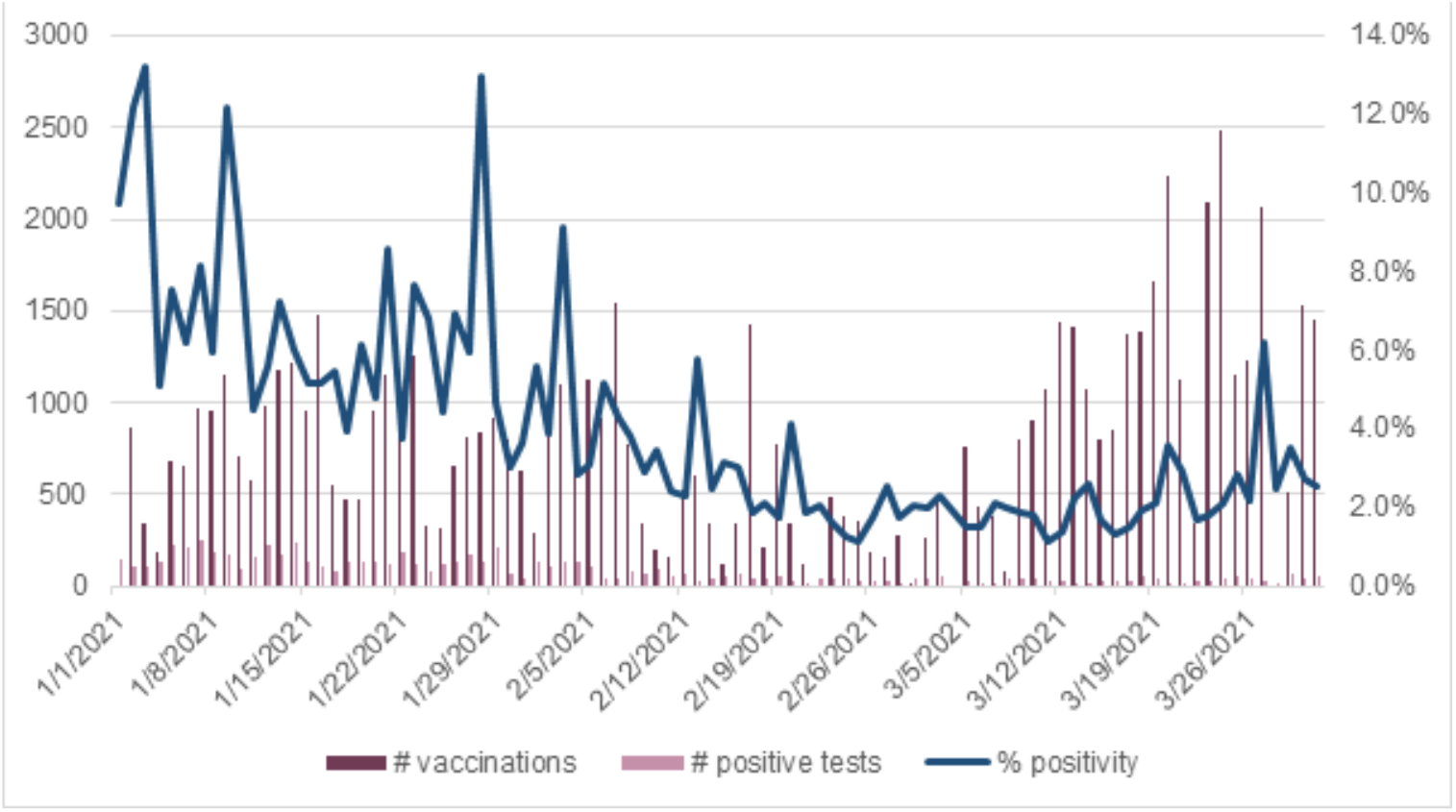
COVID-19 vaccinations and test positivity in Alachua County, Florida, from January 1^st^ – March 31^st^, 2021.

**Figure S2.**
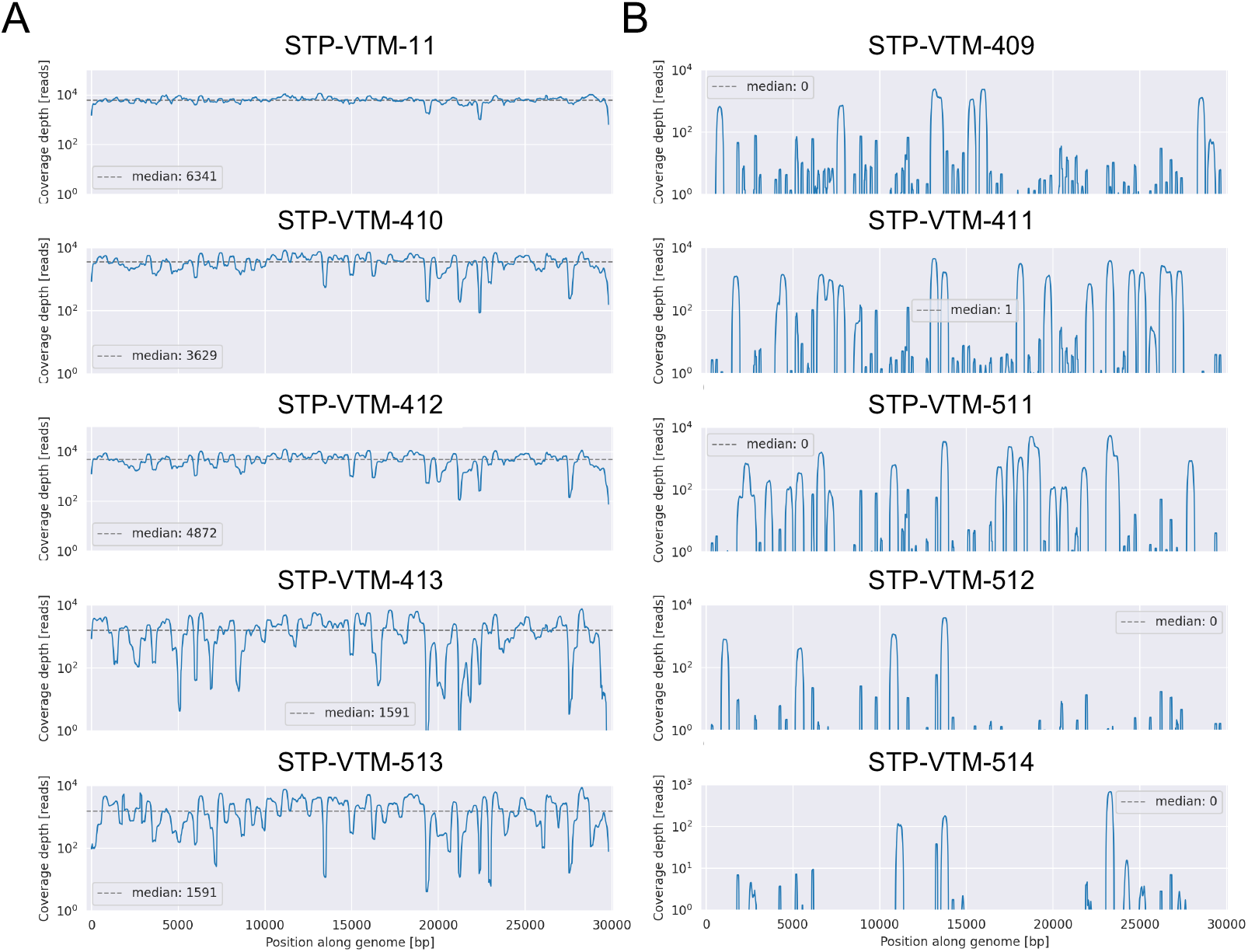
SARS-CoV-2 genome coverage for (A) five successfully and (B) five unsuccessfully sequenced vaccinated individuals. Coverage in this plot refers to the number of reads representing each position within the MN908947 reference SARS-CoV-2 genome and was provided by Illumina.

**Figure S3.**
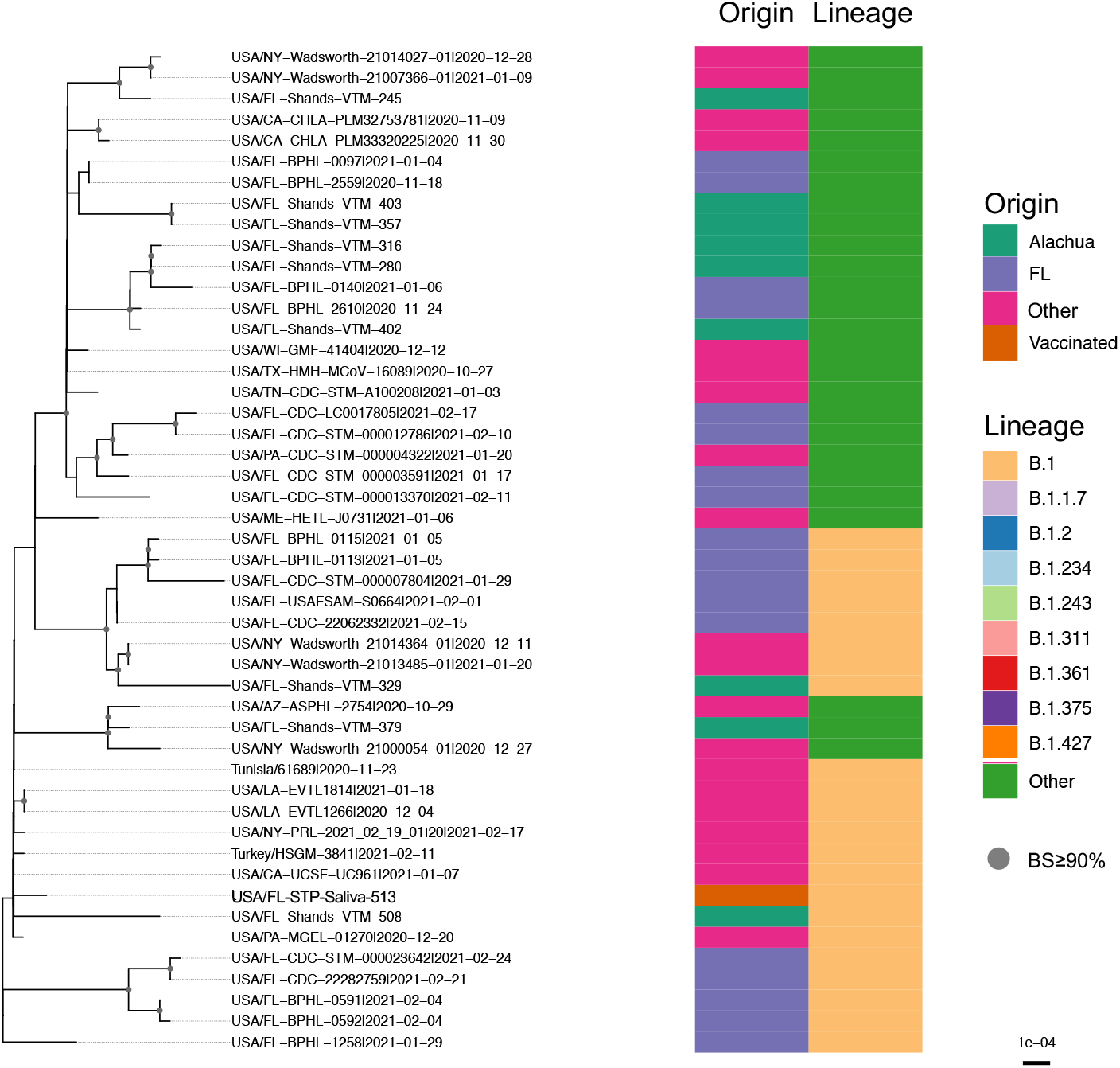
Distribution of identified lineages and geographical origin within a fragment of the phylogenetic tree containing FL-Shands-VTM-513 (B.1 lineage). Branches shown are scaled in genetic substitutions/site. Nodes with ≥90% support using bootstrap sampling are indicated by grey dots. This clade is not well-supported but represents a fragment of the tree in Figure 1 three nodes back from the FL-STP-Saliva-513 taxon. Other lineages represent lineages comprising <1% of the total sample population, though in this subtree only B.1.349 and B.1.363 are present.

